# Depressive Symptoms Are Associated With Reduced Unsupervised Training Participation in Inpatients With Subacute Stroke: A Secondary Data Analysis Study

**DOI:** 10.1101/2024.06.21.24309324

**Authors:** Kazuaki Oyake, Kaori Takahashi, Aiko Arikawa, Honoka Abe, Kunitsugu Kondo, Yohei Otaka, Satoshi Tanaka

## Abstract

**Objective:** To investigate the association between depressive symptoms and time spent in unsupervised training among inpatients with subacute stroke.

**Design:** This study was a secondary analysis of an unpublished dataset from 34 inpatients with subacute stroke (19 males; median age 65 [interquartile range, 55–75] years). Primary outcome was the median time spent in unsupervised training across three leg cycle sessions. Secondary outcomes included the Functional Independence Measure motor scores at discharge and the length of stay. Depressive symptoms were defined as the Japanese version of the Geriatric Depression Scale Short Form score of ≥7.

**Results:** Twelve participants (35.3%) had depressive symptoms. The median total time spent in unsupervised training was significantly lower in the group with depressive symptoms (367 [249–799] sec) than in the group without depressive symptoms (888 [579–901] sec), with a medium effect size (U = 57, p = 0.006, Cohen’s r = 0.46). No significant differences were found in the secondary outcomes (p > 0.05).

**Conclusions:** Depressive symptoms were associated with reduced participation in unsupervised training among inpatients after stroke. The findings highlight the importance of considering psychological factors in designing and implementing self-rehabilitation programs at the early stages of rehabilitation.

What is Known

- Meta-analysis shows that self-rehabilitation programs are as effective as conventional therapy in improving post-stroke motor function and activity, receiving their capacity to increase rehabilitation opportunities and outcomes.
- Depressive symptoms decrease physical activity and active participation in rehabilitation among individuals with stroke.

What is New

- Depressive symptoms were associated with decreased time spent in unsupervised rehabilitation among inpatients with subacute stroke.
- The findings highlight the importance of assessing and managing depressive symptoms from the early stages of rehabilitation in order to optimize patient adherence to self-rehabilitation programs.

## INTRODUCTION

Stroke is a leading cause of disability worldwide, and the increasing incidence of stroke has led to a growing demand for effective and extended rehabilitation. Self-rehabilitation programs, in which patients engage in training tasks independently without the presence of a clinician for most of the time, have emerged as a promising approach to increase rehabilitation intensity and duration while reducing the burden on healthcare providers.^1^ These programs can be delivered in various settings, including rehabilitation hospitals, physical therapy clinics, and patients’ homes, and have been shown to be as effective as conventional therapy in enhancing motor function and activity following stroke.^1^ However, poor adherence to unsupervised training can negatively impact the outcomes of self-rehabilitation programs. Therefore, it is crucial to identify potential factors that affect patient adherence to unsupervised training to optimize the effectiveness of these programs.

Depression, a common comorbidity affecting approximately one-third of stroke survivors, have shown to have a negative impact of post-stroke physical activity.^2,3^ Several quantitative and qualitative studies have reported that depressive symptoms are associated with low levels of physical activity and limited active participation in rehabilitation among inpatients with stroke.^4-6^ However, to the best of our knowledge, no study has distinguished between supervised and unsupervised training when evaluating physical activity levels and participation in rehabilitation. Consequently, the relationship between depressive symptoms and participation in unsupervised training has not been investigated in inpatients with stroke, despite the potential importance of self-rehabilitation programs in the early stages of stroke.

To address this issue, this study aimed to investigate whether depressive symptoms are associated with reduced participation in unsupervised training among inpatients with subacute stroke. We utilized an unpublished dataset from an experiment that objectively measured the time inpatients with subacute stroke spent engaged in unsupervised leg cycling training and collected several psychological measures, such as a depression scale. This unique experimental design allowed us to examine the relationship between depressive symptoms and the time spent in unsupervised training. Based on previous findings suggesting a negative impact of depressive symptoms on physical activity and rehabilitation participation,^2,3,5,6^ we hypothesized that participants with depressive symptoms would spend less time engaged in unsupervised training compared to those without depressive symptoms.

## METHODS

### Study Design

This secondary analysis study utilized an unpublished dataset obtained from a previous study conducted by our research group. The protocol of the original study was pre-registered in the University Hospital Medical Information Network (registration numbers: UMIN000036407 and UMIN000044226). The study was approved by the ethics committees of the Tokyo Bay Rehabilitation Hospital (approval number: 166-3) and Shinshu University (approval number: 5065). All participants provided written informed consent before enrollment in the study. The study was conducted in accordance with the Declaration of Helsinki of 1964, as revised in 2013.

### Participants

This study included 34 patients with stroke admitted to a convalescent rehabilitation ward in Japan. The inclusion criteria were as follows: (1) age between 40 and 80 years; (2) diagnosed with cerebral infarction or intracerebral hemorrhage; (3) at least 1 month post-stroke; (4) ability to safely perform leg cycling training; (5) currently undergoing leg cycling training; and (6) at least 1 week after admission. Patients were excluded if they met any of the following criteria: (1) limited range of motion and/or pain that could affect leg cycle performance; (2) moderate to severe dementia, as defined by a Mini-Mental State Examination (MMSE) score of <21;^7,8^ (3) unstable medical conditions, such as unstable angina, uncontrolled hypertension, or tachycardia; (4) any comorbid neurological disorders; (5) psychiatric disorders that could impede study cooperation; or (6) deemed unsuitable for participation by a physician. It should be noted that the sample size and criteria were not optimized for this secondary analysis, as the original study protocol was designed for a different purpose.

### Procedure

After obtaining informed consent, demographic and clinical data, such as age and type of stroke, were collected from patient medical records. Before evaluating participation in unsupervised training, we assessed depressive symptoms, apathy, motivation for rehabilitation, and functional outcomes. Functional outcomes included Stroke Impairment Assessment Set (SIAS) motor items,^9^ Functional Ambulation Category (FAC),^10^ Modified Rankin Scale (mRS),^11^ and Functional Independence Measure (FIM) motor and cognition items.^12^ The evaluation of unsupervised training participation consisted of three leg cycle training sessions, each conducted on separate days. The aforementioned data collection was completed within one week of the start of the procedure.

As depression has been shown to negatively affect functional recovery after stroke by limiting participation in rehabilitation,^2^ we collected additional data on the FIM motor scores at discharge and the length of stay from patient medical records after the participants were discharged from the hospital. This allowed us to examine the relationship between depressive symptoms and stroke recovery outcomes.

#### Assessments of depressive symptoms, apathy, and motivation for rehabilitation

The Geriatric Depression Scale Short Form (GDS-S) is a widely used questionnaire for screening depressive symptoms.^13^ The GDS-S also reportedly shows good sensitivity and specificity for the detection of depressive symptoms in young and middle-aged adults.^14^ Therefore, we used the Japanese version of the GDS-S, with scores of 7 or higher indicating the presence of depressive symptoms.^15^ Apathy was evaluated using the Japanese version of the Apathy Scale, with scores of 16 points or more considered indicative of apathy.^16,17^ Participants’ motivation for rehabilitation was subjectively rated using a visual analog scale (VAS).^18^ The possible score ranged from 0 (no motivation) to 100 (highest motivation), measured in millimeters on a 100-mm vertical line using a pen.

#### Assessment of the unsupervised training participation

Leg cycle training was performed on a cycle ergometer in the rehabilitation room. The physical therapist responsible for the assessment selected the appropriate cycle ergometer for each participant from the following options: Aero Bike ai-ex (Combi Wellness, Tokyo, Japan), Aero Bike 2100R (Combi Wellness, Tokyo, Japan), or Kardiomed d-cycle (Proxomed, Alzenau, Germany). The workload was set at an intensity level that each participant perceived as “somewhat hard.”^19^ The equipment used and the workload set in the first session were kept consistent for the subsequent two sessions. Participants were instructed to maintain a cycling cadence at which they could comfortably perform the training at a given workload.

The leg cycle training consisted of an initial 5-minute supervised phase, followed by an unsupervised phase lasting up to 15 minutes. During the supervised phase, physical therapists observed the participants, conversing with and encouraging them as appropriate. At the end of the supervised phase, heart rate was measured using a pulse oximeter (Oxypal mini; Nihon Kohden Co., Ltd, Tokyo, Japan) to assess the relative intensity of the supervised training. The relative training intensity was calculated as the percentage of age-predicted maximal heart rate (220 minus age).^20^ The median value of the relative training intensity across the three sessions was used for statistical analysis.

Following the completion of the supervised phase, participants transitioned directly to the unsupervised phase without a break. However, during the unsupervised phase, they were permitted to take breaks or terminate their unsupervised training at their discretion. Physical therapists monitored the participants from an area not visible to them to ensure their safety. The unsupervised phase was terminated when the participant requested to stop the training or when 15 minutes elapsed from the start of the unsupervised phase. The total time spent in unsupervised training (in seconds) was recorded as a measure of unsupervised training participation. The median value of the total time spent in unsupervised training across the three leg cycle sessions was calculated as the representative value for each participant.

Although the participants were informed about the study contents beforehand, they were not aware that the total time spent in unsupervised training was the outcome measure, making this a single-blind study design. After completing all parts of the experiment, participants received a debriefing.

### Statistical Analysis

The primary outcome was the total time spent in unsupervised training. The secondary outcomes included the FIM motor scores at discharge and the length of stay. To examine the associations of depressive symptoms with the primary and secondary outcomes, we used the Mann-Whitney U test and calculated Cohen’s r as a measure of the effect size: small, 0.10 ≤ r < 0.30; medium, 0.30 ≤ r < 0.50; large, r ≥ 0.50.^21^ As depressive symptoms have overlapping symptoms with apathy, such as loss of pleasure, reduced energy, and physical/mental slowing,^22^ we also compared the primary outcome between the groups with and without apathy. To identify confounding variables, we examined the associations of depressive symptoms with the demographic and clinical data, apathy, motivation for rehabilitation, and the relative intensity of the supervised training using the Mann-Whitney U test for continuous variables and Fisher’s exact test for categorical variables. Statistical analyses were performed using SPSS software version 28.0 (IBM Corp., NY, USA). P values < 0.05 were considered statistically significant.

## RESULTS

### Participants

Thirty-four patients with stroke were included in the analysis (Table 1). The median [interquartile range] time since stroke onset in overall participants was 91 [63–111] days. According to the GDS-S scores, 12 participants (35.3%) had depressive symptoms. The GDS-S score in the group with depressive symptoms was 8 [7–11] points, whereas that of the group without depressive symptoms was 3 [1–4] points.

**Table 1.** Demographic and clinical data, apathy, motivation for rehabilitation, and functional outcomes.

| Variable | Overall<br>(n = 34) | With depressive<br>symptoms (n = 12) | Without depressive<br>symptoms (n = 22) | p-value |
| --- | --- | --- | --- | --- |
| Age (age) | 65 [55–75] | 61 [51–75] | 67 [56–77] | 0.378 |
| Sex, female/male | 15/19 | 6/6 | 9/13 | 0.724 |
| Type of stroke,<br>ischemic/hemorrhage | 14/20 | 4/8 | 10/12 | 0.717 |
| Side of motor paresis,<br>left/right | 21/13 | 8/4 | 13/9 | 0.727 |
| Time since stroke onset<br>(days) | 91 [63–111] | 86 [76–109] | 96 [60–117] | 0.825 |
| MMSE score (points) | 27 [25–29] | 27 [25–29] | 28 [24–29] | 0.597 |
| Apathy Scale score<br>(points) | 13 [10–16] | 16 [14–18] | 11 [6–14] | 0.006 |
| Apathy Scale score ≥ 16 | 10 | 6 | 4 | 0.112 |
| VAS score for motivation<br>(mm) | 74 [87–97] | 82 [53–90] | 88 [79–97] | 0.358 |
| SIAS motor score (points) | 27 [25–29] | 22 [14–25] | 22 [17–25] | 0.849 |
| FAC score (points) | 4 [4–5] | 4 [3–5] | 4 [4–5] | 0.631 |
| mRS score (points) | 2 [1–3] | 2 [0–4] | 2 [1–3] | 0.318 |
| FIM motor score (points) | 59 [41–71] | 64 [38–67] | 57 [43–80] | 0.703 |
| FIM cognition score<br>(points) | 29 [24–31] | 29 [21–30] | 29 [24–32] | 0.339 |

Values are presented as median [interquartile range] for continuous variables and as number for categorical variables.
All p values indicate comparisons between the groups with and without depressive symptoms.
Abbreviations: MMSE, Mini-Mental State Examination; SIAS, Stroke Impairment Assessment Set; FAC, Functional Ambulation Category; mRS, modified Rankin Scale; FIM, Functional Independence Measure; VAS, Visual Analog Scale.

The Apathy Scale scores were significantly higher in the group with depressive symptoms than in the group without depressive symptoms (U = 57, p = 0.006). A total of 10 participants (29.4%) exhibited apathy, of whom 6 were classified in the group with depressive symptoms. However, there was no significant difference in the prevalence of apathy between the groups with and without depressive symptoms (p = 0.112). In addition, depressive symptoms were not significantly associated with demographic and clinical data, VAS scores for motivation, and functional outcomes (p > 0.05).

### Assessment of unsupervised training participation

No significant adverse events occurred during or after the training sessions. The median relative intensity was 61.3 [53.3–64.9]% of the age-predicted maximal heart rate in the group with depressive symptoms and 57.3 [53.2–62.3]% of the age-predicted maximal heart rate in the group without depressive symptoms. These results indicate that both groups did not exceed the recommended intensity for aerobic exercise after stroke.^19^ The relative intensity during the supervised training was not significantly different between the two groups (U = 115, p = 0.552).

The total time spent in unsupervised training was significantly shorter in the group with depressive symptoms (367 [249-799] sec) than in the group without depressive symptoms (888 [579-901] sec), with a medium effect size (U = 57, p = 0.006, Cohen’s r = 0.46; Figure 1A). However, there was no significant difference in the total time spent in unsupervised training between the 10 participants with apathy (455 [251-885] sec) and the 24 without apathy (731 [425–902] sec), despite a medium effect size (U = 74, p = 0.059, Cohen’s r = 0.32; Figure 1B).

**Figure 1.**
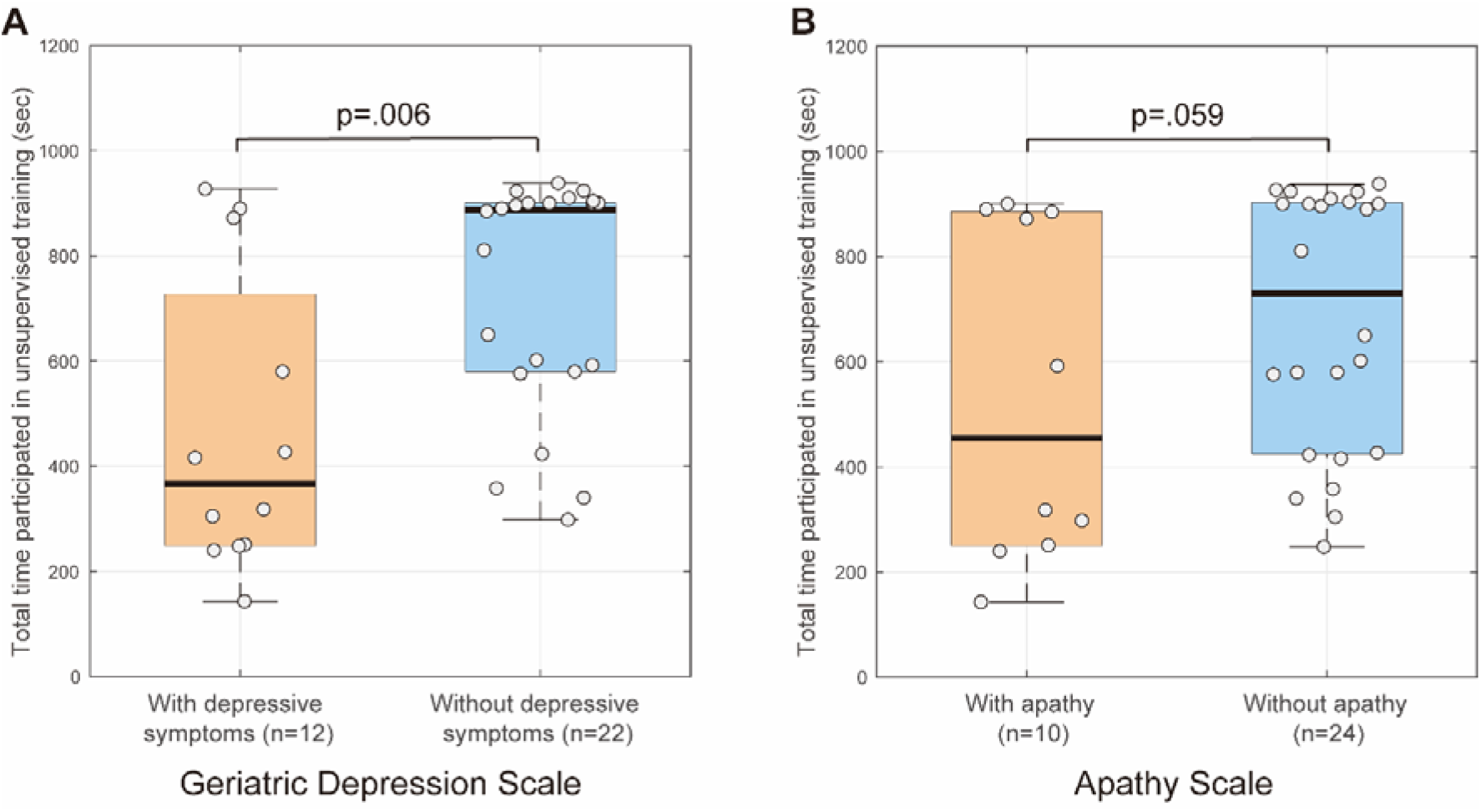
Comparisons of the total time spent in unsupervised training (A) between the groups with and without depressive symptoms and (B) between the groups with and without apathy. Data are presented as median (bold black line) and interquartile range with individual plot (grey circle).

### Secondary outcomes: FIM motor scores at discharge and length of stay

The FIM motor scores at discharge in the group with depressive symptoms were 85 [80-89] points, while those in the group without depressive symptoms were 87 [85-90] points. The median length of stay was 97 [52–115] days for the group with depressive symptoms and 101 [45–139] days for the group without depressive symptoms. There were no significant differences between the two groups for the FIM motor scores at discharge (U = 103, p = 0.303, Cohen’s r = 0.18) and the length of stay (U = 119, p = 0.638, Cohen’s r = 0.08).

## DISCUSSION

In this study, we investigated the association between depressive symptoms and the total time spent in unsupervised training among inpatients with subacute stroke. We observed that 35.3% of participants had depressive symptoms. Given that the median time since stroke onset was approximately 3 months (91 [63–111] days), the prevalence of depressive symptoms in this study was similar to data from a previous study indicating that the prevalence of depressive symptoms was 35.3% at 3 months after stroke.^23^ We found that shorter time spent in unsupervised training in the group with depressive symptoms than in the group without depressive symptoms, suggesting that depressive symptoms are associated with reduced unsupervised training participation in inpatients with subacute stroke.

Depressive symptoms have been reported to have a negative effect on active participation in rehabilitation among hospitalized patients with stroke.^4-6^ However, these studies primarily focused on overall physical activity and participation, leaving the specific influence of depressive symptoms on unsupervised training participation unclear. To our knowledge, this is the first study to specifically investigate the relationship between depressive symptoms and unsupervised training participation in a hospital setting, which is an important strength of this study. Given that poor adherence to unsupervised training may affect self-rehabilitation outcomes,^1^ the results of this study highlight the need for considering psychological factors in the early stages of rehabilitation.

Several factors may contribute to the observed relationship between depressive symptoms and reduced time spent in unsupervised training. Importantly, both groups performed supervised training at an intensity level that did not exceed the guidelines’ recommendations,^19^ with no significant difference between the groups. These results suggest that the difference in unsupervised training time is unlikely to be attributed to variations in the training intensity during the experiment. One possibility is that the core features of depressive symptoms, such as loss of interest/pleasure, feeling down/hopeless, fatigue, and changes in appetite, may hinder patients’ engagement in unsupervised training.^24^ This idea is partially supported by the association between apathy and the time spent in unsupervised training (Cohen’s r = 0.32) observed in this study, although the difference did not reach statistical significance (p = 0.059). However, it is important to note that our study did not directly investigate these mechanisms, and further research is needed to examine which specific depressive symptoms influence unsupervised training participation in patients with stroke.

As depression after stroke have been known to affect functional recovery as well as participation in rehabilitation,^2,25^ we expected that participants with depressive symptoms would show lower FIM motor scores at discharge and/or a longer length of stay, as well as a shorter time spent in unsupervised training than those without depressive symptoms. However, there were no significant differences in the FIM motor scores at discharge and the length of stay between the groups with and without depressive symptoms. This discrepancy may be attributed to three reasons. First, participants who were considered to have depressive symptoms according to the GDS-S scores were not diagnosed with depression by the psychiatrists. Second, the relationship between depressive symptoms and functional recovery may be more complex than initially assumed. It is possible that other factors, such as the quality and intensity of supervised rehabilitation or individual differences in functional recovery, may have lessened the impact of depressive symptoms on functional recovery.^2,26^ Third, the relatively short duration of the study may not have been sufficient to capture the long-term effects of depressive symptoms on functional recovery.

The present findings suggest that it is crucial for healthcare providers to prioritize the early identification of stroke patients with depressive symptoms and provide appropriate mental health support to optimize their engagement in self-rehabilitation. Thus, this study emphasizes the significance of a multidisciplinary-team approach in stroke rehabilitation, which should include not only physical and occupational therapists but also mental health professionals such as psychologists and psychiatrists.^2,25^ In addition, it is important to implement psychological strategies, such as empathic and motivational communications, to enhance their adherence to unsupervised training and optimize the outcomes of self-rehabilitation programs in inpatients with depressive symptoms after stroke.^6,25,27,28^

### Study Limitation

Our study has several limitations. First, as this was a secondary analysis using an unpublished dataset, the findings should be considered preliminary. Future studies with *a priori* hypotheses and study designs tailored to investigate the relationship between depressive symptoms and unsupervised training participation are needed to confirm our results. Second, the sample size was not optimized for the current study’s objectives, as the original study was designed for a different purpose. Third, we did not investigate the long-term effects of depressive symptoms on unsupervised training participation or the impact on functional recovery. Longitudinal studies with larger sample sizes are warranted to address these questions. Finally, future research should explore the effectiveness of interventions designed to mitigate the impact of depressive symptoms on unsupervised training participation, such as motivational strategies, cognitive behavioral therapies, or socio-psychological support programs.^6,25,27-30^

## CONCLUSION

The results suggest that depressive symptoms may significantly reduce unsupervised training time in subacute stroke inpatients. These findings highlight the importance of screening for depressive symptoms and providing appropriate mental health support to optimize patient adherence to self-rehabilitation programs in stroke rehabilitation.

## Data Availability

All data produced in the present study are available upon reasonable request to the authors.

## Data Availability statement

Data are available in the supplementary file.

## Funding or grants or equipment provided for the project from any source

This work was supported by a grant from JSPS KAKENHI Grant Number JP20K21752 to S.T.

## Financial benefits to the authors

None.

## Details of any previous presentation of the research, manuscript, or abstract in any form

None.

## Acknowledgments

The authors acknowledge the use of Claude 3 Opus (Anthropic, San Francisco, CA, USA) for generating preliminary drafts and English editing assistance. Final manuscript revisions were done by the human researchers.

## REFERENCES

1. Everard G, Luc A, Doumas I, et al. Self-rehabilitation for post-stroke motor function and activity: a systematic review and meta-analysis. Neurorehabil Neural Repair. 2021;35:1043–1058. doi:10.1177/15459683211048773

2. Towfighi A, Ovbiagele B, El Husseini N, et al. Poststroke depression: a scientific statement for healthcare professionals from the American Heart Association/American Stroke Association. Stroke. 2017;48:e30–e43. doi:10.1161/str.0000000000000113

3. Thilarajah S, Mentiplay BF, Bower KJ, et al. Factors associated with post-stroke physical activity: a systematic review and meta-analysis. Arch Phys Med Rehabil. 2018;99:1876–1889. doi:10.1016/j.apmr.2017.09.117

4. Kunkel D, Fitton C, Burnett M, Ashburn A. Physical inactivity post-stroke: a 3-year longitudinal study. Disabil Rehabil. 2015;37:304–10. doi:10.3109/09638288.2014.918190

5. Skidmore ER, Whyte EM, Holm MB, et al. Cognitive and affective predictors of rehabilitation participation after stroke. Arch Phys Med Rehabil. 2010;91:203–7. doi:10.1016/j.apmr.2009.10.026

6. Oyake K, Sue K, Sumiya M, Tanaka S. Physical therapists use different motivational strategies for stroke rehabilitation tailored to an individual’s condition: a qualitative study. Phys Ther. 2023;103: pzad034. doi:10.1093/ptj/pzad034

7. Folstein MF, Folstein SE, McHugh PR. “Mini-mental state”. A practical method for grading the cognitive state of patients for the clinician. J Psychiatr Res. 1975;12:189–98.

8. Perneczky R, Wagenpfeil S, Komossa K, Grimmer T, Diehl J, Kurz A. Mapping scores onto stages: mini-mental state examination and clinical dementia rating. Am J Geriatr Psychiatry. 2006;14:139–44. doi:10.1097/01.JGP.0000192478.82189.a8

9. Chino N, Sonoda S, Domen K, Saitoh E, Kimura A. Stroke Impairment Assessment Set (SIAS) A new evaluation instrument for stroke patients. Jpn J Rehabil Med. 1994;31:119–125. doi:10.2490/jjrm1963.31.119

10. Holden MK, Gill KM, Magliozzi MR, Nathan J, Piehl-Baker L. Clinical gait assessment in the neurologically impaired. Reliability and meaningfulness. Phys Ther. 1984;64:35–40. doi:10.1093/ptj/64.1.35

11. van Swieten JC, Koudstaal PJ, Visser MC, Schouten HJ, van Gijn J. Interobserver agreement for the assessment of handicap in stroke patients. Stroke. 1988;19:604–7. doi:10.1161/01.str.19.5.604

12. Keith RA, Granger CV, Hamilton BB, Sherwin FS. The functional independence measure: a new tool for rehabilitation. Adv Clin Rehabil. 1987;1:6–18.

13. Sheikh JI. Geriatric depression scale (GDS). Recent evidence and development of a shorter version. Clin Gerontol 1986;5: 165–172.

14. Guerin JM, Copersino ML, Schretlen DJ. Clinical utility of the 15-item geriatric depression scale (GDS-15) for use with young and middle-aged adults. J Affect Disord. 2018;241:59–62. doi:10.1016/j.jad.2018.07.038

15. Sugishita K, Sugishita M, Hemmi I, Asada T, Tanigawa T. A Validity and Reliability Study of the Japanese Version of the Geriatric Depression Scale 15 (GDS-15-J). Clin Gerontol. 2017;40:233–240. doi:10.1080/07317115.2016.1199452

16. Starkstein SE, Fedoroff JP, Price TR, Leiguarda R, Robinson RG. Apathy following cerebrovascular lesions. Stroke. 1993;24(11):1625–30. doi:10.1161/01.str.24.11.1625

17. Okada K, Kobayashi S, Yamagata S, Takahashi K, Yamaguchi S. Poststroke apathy and regional cerebral blood flow. Stroke. 1997;28:2437–41. doi:10.1161/01.str.28.12.2437

18. Tanaka M, Ishii A, Watanabe Y. Neural mechanism of facilitation system during physical fatigue. PLoS One. 2013;8:e80731. doi:10.1371/journal.pone.0080731

19. Billinger SA, Arena R, Bernhardt J, et al. Physical activity and exercise recommendations for stroke survivors: a statement for healthcare professionals from the American Heart Association/American Stroke Association. Stroke. 2014;45:2532–53. doi:10.1161/STR.0000000000000022

20. Fletcher GF, Ades PA, Kligfield P, et al. Exercise standards for testing and training: a scientific statement from the American Heart Association. Circulation. 2013;128:873–934. doi:10.1161/CIR.0b013e31829b5b44

21. Fritz CO, Morris PE, Richler JJ. Effect size estimates: current use, calculations, and interpretation. J Exp Psychol Gen. 2012;141:2–18. doi:10.1037/a0024338

22. Tay J, Morris RG, Markus HS. Apathy after stroke: diagnosis, mechanisms, consequences, and treatment. Int J Stroke. 2021;16:510–518. doi:10.1177/1747493021990906

23. Dong L, Williams LS, Brown DL, Case E, Morgenstern LB, Lisabeth LD. Prevalence and course of depression during the first year after mild to moderate stroke. J Am Heart Assoc. 2021;10:e020494. doi:10.1161/jaha.120.020494

24. Soini E, Rosenström T, Määttänen I, Jokela M. Physical activity and specific symptoms of depression: a pooled analysis of six cohort studies. J Affect Disord. 2024;348:44–53. doi:10.1016/j.jad.2023.12.039

25. Winstein CJ, Stein J, Arena R, et al. Guidelines for adult stroke rehabilitation and recovery: a guideline for healthcare professionals from the American Heart Association/American Stroke Association. Stroke. 2016;47:e98–e169. doi:10.1161/STR.0000000000000098

26. Boyd LA, Hayward KS, Ward NS, et al. Biomarkers of stroke recovery: consensus-based core recommendations from the Stroke Recovery and Rehabilitation Roundtable. Int J Stroke. 2017;12:480–493. doi:10.1177/1747493017714176

27. Oyake K, Yamauchi K, Inoue S, et al. A multicenter explanatory survey of patients’ and clinicians’ perceptions of motivational factors in rehabilitation. Commun Med (Lond). 2023;3:78. doi:10.1038/s43856-023-00308-7

28. yake K, Watanabe S, Takeuchi A, et al. Applying a motivational instructional design model to stroke rehabilitation: a feasibility study on occupational and swallowing therapies. Arch Rehabil Res Clin Transl. 2024:100344. doi:10.1016/j.arrct.2024.100344

29. McGrane N, Galvin R, Cusack T, Stokes E. Addition of motivational interventions to exercise and traditional physiotherapy: a review and meta-analysis. Physiotherapy. 2015;101:1–12. doi:10.1016/j.physio.2014.04.009

30. Oyake K, Suzuki M, Otaka Y, Momose K, Tanaka S. Motivational strategies for stroke rehabilitation: a Delphi study. Arch Phys Med Rehabil. 2020;101:1929–1936. doi:10.1016/j.apmr.2020.06.007

